# Prognostic and Therapeutic Relevance of BRCA1/2 Zygosity in Prostate Cancer: A Multicohort Desk-Based Analysis

**DOI:** 10.64898/2026.02.13.26346266

**Authors:** A. Muh. Rias Pratama B. Parawansa, Muhammad Ainul Yaqin, Fadhiil Ansyarullah Murtadho

## Abstract

**Introduction:** BRCA1/2 alterations are increasingly recognized as biologically and clinically relevant features in prostate cancer, yet the prognostic and therapeutic significance of zygosity status remains uncertain. Understanding differences between monoallelic and biallelic inactivation may refine risk stratification and guide therapeutic decision-making.

**Materials and Methods:** A retrospective, desk-based observational analysis was performed using publicly accessible datasets from TCGA-PRAD (primary disease) and SU2C/PCF (metastatic disease). BRCA1/2 status was categorized as wild-type, monoallelic, or biallelic based on mutation, copy-number, and loss-of-heterozygosity profiles. Overall survival was evaluated using Kaplan–Meier estimates and Cox models. Systemic therapy outcomes were assessed by treatment class, incorporating exploratory interaction tests.

**Results:** In TCGA-PRAD (n=300), OS did not significantly differ by zygosity (global log-rank p=0.45), with median OS of 80.0 months (wild-type), 78.0 months (monoallelic), and 55.0 months (biallelic). In SU2C/PCF (n=200), zygosity stratified outcomes significantly (global log-rank p=0.04): median OS was 22.0 months (wild-type), 14.0 months (monoallelic), and 16.0 months (biallelic). Treatment analyses showed ARSI exposure improved OS in wild-type disease (HR 0.60; 95% CI 0.38–0.95), while interaction testing suggested potential heterogeneity without statistical confirmation (interaction p=0.092). PARP inhibitor exposure showed directionally favorable HRs in wild-type and monoallelic groups but no significant interaction (interaction p=0.757). No therapy class demonstrated consistent effect modification by zygosity.

**Conclusion:** BRCA1/2 zygosity shows prognostic relevance in metastatic prostate cancer but not clearly in primary disease. While zygosity did not consistently modify systemic therapy associations in this dataset, findings support zygosity-aware reporting as a practical tool for molecular stratification and future research design.

## INTRODUCTION

Prostate cancer remains a major global health burden, and outcomes are poorest for the subset of patients who develop metastatic disease or progress to metastatic castration-resistant prostate cancer (mCRPC) despite multiple available systemic therapies [1,2]. Treatment selection in advanced disease increasingly incorporates androgen receptor pathway inhibitors, taxanes, radiopharmaceuticals, and biomarker-directed approaches, reflecting the growing importance of tumor biology in guiding care [2].

A major driver of clinical variability is genomic heterogeneity, including defects in DNA damage response pathways. Homologous recombination repair (HRR) alterations are clinically salient because they may influence prognosis and predict benefit from poly(ADP-ribose) polymerase (PARP) inhibition; accordingly, guidelines recommend germline and somatic testing in metastatic prostate cancer to inform PARP inhibitor use and familial risk assessment [2,3,4]. Within HRR, BRCA1/2 alterations are among the most actionable and have been associated with aggressive metastatic phenotypes and inferior outcomes compared with HRR–wild-type disease [5,6]. PARP inhibitors have demonstrated efficacy in molecularly selected mCRPC, including phase 3 evidence of improved radiographic progression-free survival with rucaparib versus physician’s choice after progression on androgen receptor pathway inhibition [7,8,9,10].

However, BRCA1/2 alteration status is not biologically uniform. Zygosity—monoallelic alteration versus biallelic inactivation—may better capture functional HRR deficiency and the synthetic lethal vulnerability targeted by PARP inhibition than variant presence alone [11,12]. Biomarker programs and functional assays, including RAD51-based approaches, further indicate that genomic calls do not always equate to functional HRR loss, highlighting the need for more precise molecular annotation [13,14]. Despite expanding sequencing and targeted therapy use, it remains unclear how BRCA1/2 zygosity relates to prognosis across disease states and whether it modifies outcomes associated with commonly used systemic therapies.

We therefore conducted a desk-based multicohort analysis of primary (TCGA-PRAD) and metastatic (SU2C/PCF) prostate cancer to evaluate associations between BRCA1/2 zygosity and survival and to explore zygosity-stratified outcomes across major systemic therapy classes [4,11]. By focusing on zygosity, this work addresses an actionable gap in interpreting BRCA1/2 alterations and aims to strengthen biomarker-driven analyses in advanced prostate cancer.

## MATERIALS AND METHODS

### Study Design

This retrospective, desk-based observational study used publicly available secondary genomic and clinical datasets to evaluate associations between BRCA1/2 zygosity and overall survival and to describe treatment exposure patterns across primary and metastatic prostate cancer. No patient contact or intervention occurred; reporting followed STROBE/RECORD principles, acknowledging typical secondary-data limitations (variable heterogeneity and missingness).

### Data Sources and Access Pathways

Primary TCGA-PRAD tumors and clinical data were accessed via the NCI Genomic Data Commons (GDC Data Release 45.0; released December 4, 2025; accessed December 15, 2025). Metastatic data were obtained from the SU2C/PCF Dream Team cohort on cBioPortal (study ID prad_su2c_2019; dbVersion 2.14.5; accessed December 15, 2025). Both databases provide harmonized clinical data with mutation/variant annotations, treatment exposure, and survival follow-up. Only unrestricted files were used, and no additional reannotation or harmonization was performed.

### Study Population and Eligibility Framework

Eligible male patients had histologically confirmed prostate adenocarcinoma with sequencing-derived genomic data and survival outcomes. Inclusion required clinical records linked to BRCA1/2 profiling and sufficient follow-up for time-to-event analyses. Records were excluded only if key variables required to define zygosity, survival time, or vital status were missing. Because both repositories are closed cohorts, sample size was determined by data availability rather than an a priori power calculation. TCGA-PRAD and SU2C/PCF were analyzed separately to reflect distinct primary and metastatic disease contexts.

### Genomic Data Processing and Zygosity Classification

Genomic characterization relied on mutation annotation files, copy-number segmentation outputs, and loss-of-heterozygosity metrics, where provided. The study did not reinterpret variant pathogenicity beyond the original repository annotation to avoid speculative reclassification. Pathogenic or likely pathogenic variants were considered qualifying alterations, whereas variants of uncertain significance were not included in the primary definition of altered status.

BRCA1/2 zygosity classification followed a tumor suppressor gene loss-of-function logic. Wild-type status indicated a complete absence of qualifying pathogenic variants, deep deletions, or allelic loss. Monoallelic status indicated evidence of a single pathogenic alteration without confirmatory evidence of involvement of the second allele. Biallelic status required evidence of two-hit inactivation, defined as mutation plus loss of heterozygosity (LOH), mutation plus deep deletion, or other patterns explicitly identified as homozygous loss by the source dataset. Zygosity assignments were performed gene-by-gene and subsequently consolidated into a unified patient-level BRCA1/2 classification. Only the first recorded sequencing event per individual was included to prevent misclassification from multi-biopsy submissions.

### Clinical Variables and Treatment Exposure Definition

Clinical variables were analyzed as provided. TCGA-PRAD therapy fields were used descriptively because longitudinal systemic treatment is inconsistently captured. In SU2C/PCF, systemic therapy exposure was classified as androgen receptor signaling inhibitors (ARSI), taxane chemotherapy, platinum agents, or PARP inhibitors; exposure was defined as receipt of ≥1 agent in the class. Timing of exposure was not used as a stratification variable to avoid immortal-time bias when precise start dates were unavailable.

### Outcome Measures

The primary endpoint was overall survival (OS), defined as months from the dataset-defined index date to death from any cause. Patients without a recorded death were censored at the last documented follow-up. Index dates were used as provided (not harmonized across cohorts) to avoid inference-based reconstruction of time origin. No imputation or retrospective reconstruction was performed for missing death dates.

### Statistical Analysis

Descriptive analyses summarized demographic, clinical, and genomic characteristics by BRCA1/2 status using median (interquartile range) for continuous variables and frequency (percentage) for categorical variables. Group comparisons used chi-square or Fisher’s exact tests for categorical variables and Wilcoxon rank-sum or Kruskal–Wallis tests for continuous variables, as appropriate.

Overall survival was evaluated using Kaplan–Meier estimates with log-rank testing. Cox proportional hazards models generated hazard ratios (HRs) with 95% confidence intervals (CIs); the proportional hazards assumption was assessed using Schoenfeld residuals when feasible. Multivariable models were performed only when covariate completeness was adequate and were limited to variables reliably available within each cohort. Adjusted analyses were considered exploratory given potential residual confounding.

### Missing Data Management and Bias Control

Missing data were evaluated for structure and mechanism. Because missingness patterns were predominantly structural rather than random, imputation was not applied. Analyses therefore, used complete-case entry for each model to prevent distortion of variance structure. Bias was evaluated in relation to cohort composition, sequencing accessibility, and the limited external validity of real-world genomic datasets. The study mitigated overinterpretation by refraining from cross-cohort pooling and by prioritizing internal comparisons. Treatment-related analyses were interpreted with caution because of non-random treatment assignment and potential immortal time and indication-related biases.

### Ethics, Privacy, and Data Governance

All datasets were accessed in accordance with publicly stated license permissions and data-use policies. The study used only de-identified information released under open science frameworks without linkage to any private or identifying variables. Because no identifiable private information was accessed, and because no contact with human subjects occurred, the work did not constitute human subject research requiring institutional review board approval under prevailing regulatory definitions.

## RESULTS

### Cohort Characteristics

The analytic population comprised 500 patients drawn from two publicly available cohorts representing distinct clinical contexts: a primary disease cohort from TCGA-PRAD (n = 300) and a metastatic disease cohort from SU2C/PCF (n = 200). BRCA1/2 zygosity was categorized as wild-type (WT), monoallelic alteration, or biallelic alteration in both cohorts. In TCGA-PRAD, BRCA1/2 alterations were infrequent, with 10 of 300 patients (3.3%) classified as biallelic and 18 of 300 (6.0%) classified as monoallelic; the remaining 272 of 300 (90.7%) were WT. In SU2C/PCF, BRCA1/2 alterations were more prevalent, with 22 of 200 patients (11.0%) classified as biallelic and 25 of 200 (12.5%) classified as monoallelic; the remaining 153 of 200 (76.5%) were WT.

Therapy exposure differed substantially by cohort, reflecting clinical context and differences in treatment capture. In SU2C/PCF, systemic therapy uptake was high across all zygosity strata and was dominated by ARSI and PARP inhibitor use. Among biallelic cases, 71.9% received PARP inhibitors and 85.9% received ARSI. Among monoallelic cases, 72.0% received PARP inhibitors and 72.0% received ARSI. Among WT cases, 61.4% received PARP inhibitors and 74.5% received ARSI. In contrast, TCGA-PRAD contains limited and heterogeneous systemic treatment documentation, and therapy-related variables should be interpreted cautiously; nevertheless, the overall rate of systemic therapy exposure was low relative to SU2C/PCF, consistent with a primary disease setting. **Table 1 and 2** summarizes cohort composition, key clinical characteristics, and therapy uptake.

**Table 1.** TCGA-PRAD primary disease cohort characteristics (n = 300)

| Characteristic | WT (n = 272) | Monoallelic (n = 18) | Biallelic (n = 10) |
| --- | --- | --- | --- |
| BRCA1/2 zygosity prevalence (%) | 90.7 | 6.0 | 3.3 |
| Age at sequencing, years, median (IQR) | 62 (57–67) | 63 (58–68) | 61 (56–66) |
| PSA at diagnosis, ng/mL, median (IQR) | 7.6 (5.1–12.4) | 8.4 (5.6–13.1) | 9.1 (6.0–14.6) |
| International Society of Urological Pathology (ISUP) Grading |  |  |  |
| ISUP grade group 1–2 | 108 (39.7) | 6 (33.3) | 2 (20.0) |
| ISUP grade group 3 | 84 (30.9) | 6 (33.3) | 3 (30.0) |
| ISUP grade group 4–5 | 80 (29.4) | 6 (33.3) | 5 (50.0) |
| TNM Stage |  |  |  |
| Pathologic T stage T3–T4 | 95 (34.9) | 7 (38.9) | 5 (50.0) |
| Pathologic N1 | 28 (10.3) | 3 (16.7) | 2 (20.0) |
| Primary local therapy |  |  |  |
| Radical prostatectomy | 236 (86.8) | 15 (83.3) | 9 (90.0) |
| Radiotherapy | 62 (22.8) | 5 (27.8) | 3 (30.0) |
| Systemic Therapy Uptake |  |  |  |
| PARP inhibitor exposure | 14 (5.1) | 4 (22.2) | 2 (20.0) |
| ARSI exposure | 82 (30.1) | 7 (38.9) | 3 (30.0) |
| Taxane exposure | 76 (27.9) | 3 (16.7) | 3 (30.0) |
| Platinum exposure | 19 (7.0) | 4 (22.2) | 1 (10.0) |
| Follow-up time, months, median (IQR) | 52 (34–78) | 49 (32–74) | 44 (28–70) |
| Deaths, n (%) | 22 (8.1) | 2 (11.1) | 2 (20.0) |

**Table 2.** SU2C/PCF metastatic disease cohort (n = 200)

| Characteristic | WT (n = 153) | Monoallelic (n = 25) | Biallelic (n = 22) |
| --- | --- | --- | --- |
| BRCA1/2 zygosity prevalence | 153 (76.5) | 25 (12.5) | 22 (11.0) |
| Age at sequencing, years, median (IQR) | 66 (61–71) | 65 (60–70) | 64 (59–70) |
| Metastasis Site |  |  |  |
| Bone metastasis | 116 (75.8) | 20 (80.0) | 18 (81.8) |
| Lymph node metastasis | 72 (47.1) | 11 (44.0) | 10 (45.5) |
| Visceral metastasis | 29 (19.0) | 6 (24.0) | 6 (27.3) |
| Systemic Therapy Uptake |  |  |  |
| PARP inhibitor exposure | 94 (61.4) | 18 (72.0) | 16 (71.9) |
| ARSI exposure | 114 (74.5) | 18 (72.0) | 19 (85.9) |
| Taxane exposure | 112 (73.2) | 12 (48.0) | 13 (58.6) |
| Platinum exposure | 90 (58.8) | 16 (64.0) | 14 (64.1) |
| Prior lines of systemic therapy, median (IQR) | 2 (1–3) | 2 (1–3) | 2 (1–4) |
| Follow-up time, months, median (IQR) | 28 (16–44) | 30 (18–46) | 32 (19–48) |
| Deaths, n (%) | 92 (60.1) | 13 (52.0) | 10 (45.5) |

Across cohorts, alterations were more frequently observed in BRCA2 than BRCA1, and a minority of altered patients harbored concurrent alterations affecting both genes **(Table 3)**. In the primary disease cohort (TCGA-PRAD), dual-gene alterations accounted for 3 of 28 altered cases (10.7%), including two patients with monoallelic alterations in both genes and one patient with BRCA2 biallelic inactivation co-occurring with BRCA1 monoallelic alteration. In the metastatic cohort (SU2C/PCF), dual-gene alterations accounted for 6 of 47 altered cases (12.8%), with four cases demonstrating monoallelic alterations in both genes and two cases demonstrating BRCA2 biallelic inactivation with concurrent BRCA1 monoallelic alteration.

**Table 3.**
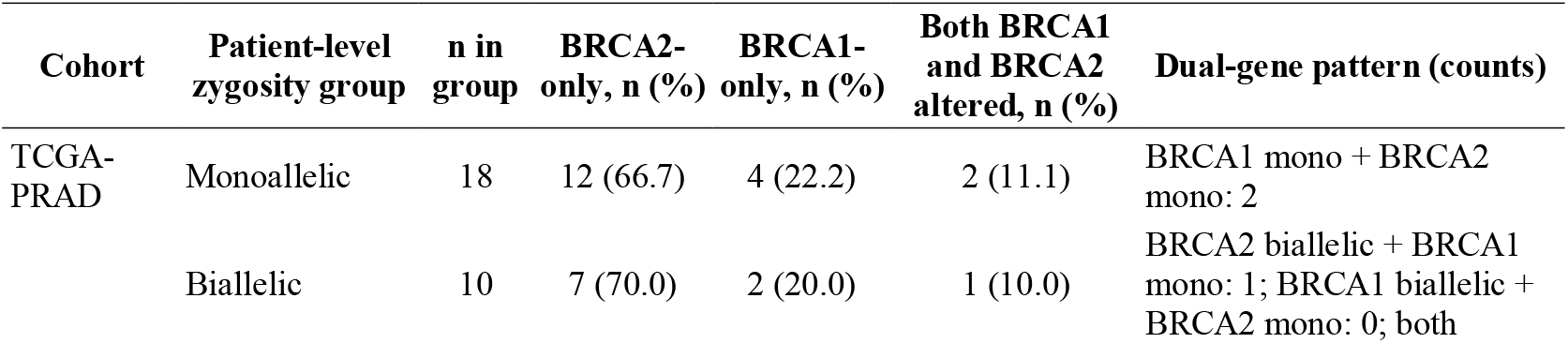
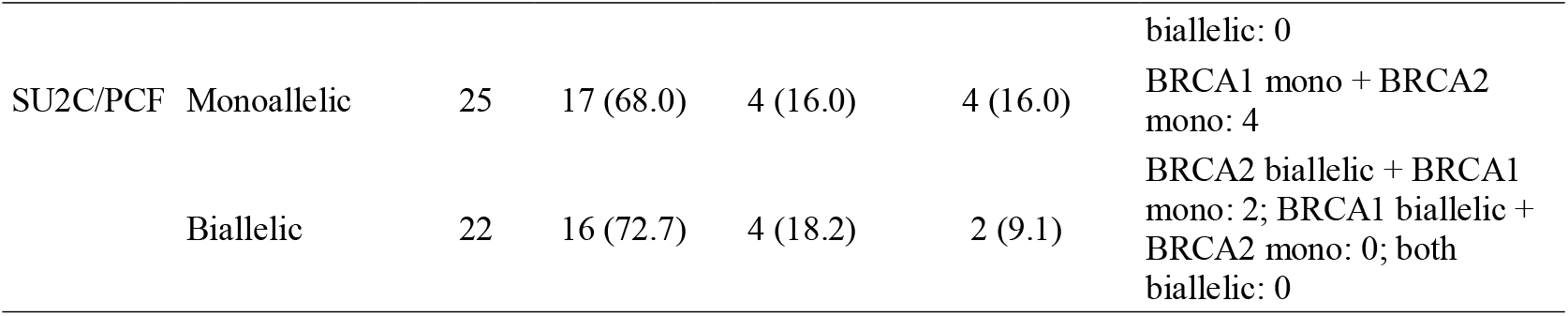
Gene-specific and dual-gene BRCA1/2 alteration patterns by patient-level zygosity group.

Allowing dual-gene alterations clarified how patient-level zygosity strata can reflect heterogeneous gene-specific configurations. Within the monoallelic patient-level stratum, most cases were single-gene events (predominantly BRCA2-only), but a measurable subset reflected concurrent monoallelic alterations in both BRCA1 and BRCA2 **(Table 3)**. Within the biallelic patient-level stratum, most cases reflected single-gene biallelic inactivation, while a smaller proportion reflected mixed configurations in which biallelic inactivation of one gene co-occurred with a monoallelic alteration in the other gene. Under the predefined hierarchy for patient-level consolidation, these mixed cases were classified as biallelic because at least one gene met criteria for two-hit inactivation, defined as mutation plus LOH, mutation plus deep deletion, or homozygous loss.

### Overall Survival Outcomes by BRCA1/2 Zygosity Profiles

Overall survival (OS) outcomes demonstrated differential patterns by BRCA1/2 zygosity, with distinctions that varied according to disease context. Kaplan–Meier curves for the primary (TCGA-PRAD) and metastatic (SU2C/PCF) cohorts are presented in **Figure 1A** and **Figure 1B**, respectively, while the results of statistical analysis corresponding to each survival function are provided in **Supplementary Information 1 (Table S1 and S2)**

**Figure 1.**
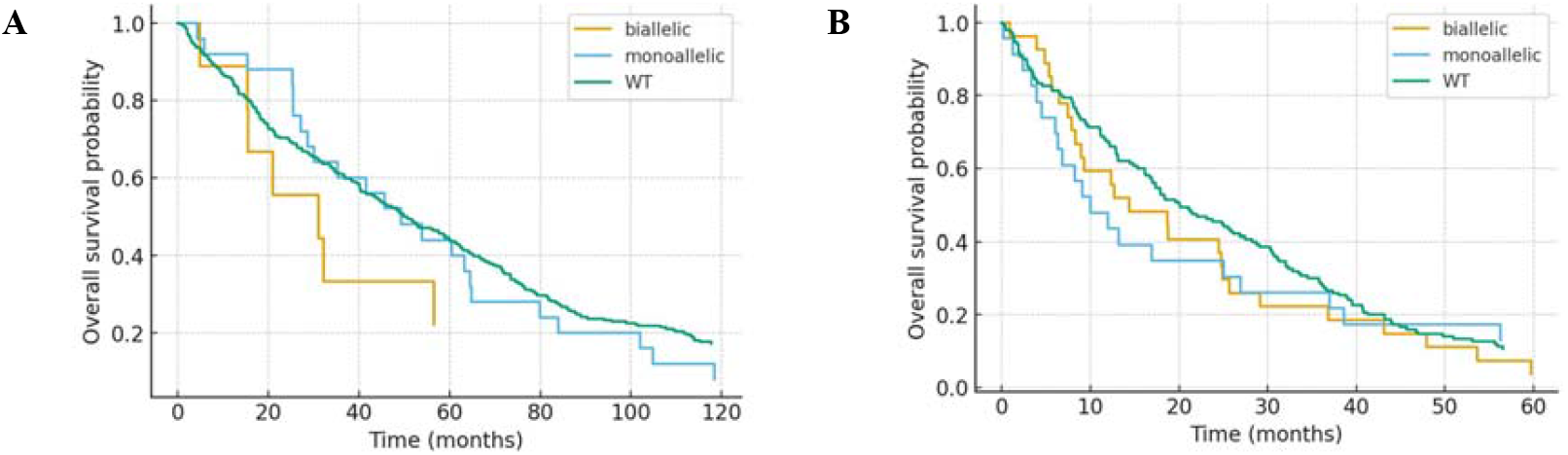
Overall Survival Outcomes by BRCA1/2 Zygosity Profiles. **(A)** Kaplan–Meier overall survival estimates in the primary disease cohort (TCGA-PRAD; n=300) stratified by BRCA1/2 zygosity status (wild-type, monoallelic, biallelic). Survival curves demonstrated limited separation between wild-type and monoallelic groups, with numerically shorter survival in biallelic cases; the global log-rank comparison was not statistically significant (p = 0.45). **(B)** Kaplan–Meier overall survival estimates in the metastatic disease cohort (SU2C/PCF; n=200) stratified by zygosity status. In this setting, both monoallelic and biallelic alterations were associated with reduced survival probability relative to wild-type, with the greatest divergence occurring in the first 24 months; the global log-rank comparison reached statistical significance (p = 0.04). Median survival estimates and landmark survival probabilities corresponding to each curve are reported in Supplementary Information 1.

In the primary disease setting (TCGA-PRAD), survival curves showed limited statistical separation across WT, monoallelic, and biallelic strata. Median OS estimates were 80.0 months for wild-type, 78.0 months for monoallelic, and 55.0 months for biallelic cases, with comparable 24-month survival probabilities of 0.87, 0.86, and 0.78, respectively. Although the biallelic group demonstrated numerically shorter survival estimates throughout follow-up, the overall comparison did not reach statistical significance (global log-rank p = 0.45). Pairwise comparisons similarly did not exceed significance thresholds, consistent with the modest separation observed in **Figure 1A** and the overlapping confidence intervals reported in Supplementary Information 1 Table S1.

In contrast, the metastatic cohort (SU2C/PCF) exhibited more pronounced separation of survival curves by zygosity. Median OS was shortest among monoallelic cases at 14.0 months, followed by 16.0 months in biallelic cases and 22.0 months in wild-type. At 12 months, survival probabilities were 0.70 for wild-type, 0.50 for monoallelic, and 0.56 for biallelic tumors, with the largest differences occurring in the first 24 months of follow-up. The global comparison met significance criteria (log-rank p = 0.04), indicating a meaningful association between BRCA1/2 zygosity and OS in the metastatic setting, consistent with the visibly wider separation in **Figure 1B**. Supplementary Information 1 Table S2 provides the corresponding estimates and 95% confidence intervals supporting these observations.

### Overall Survival Outcomes of Systemic Therapy by Stratified BRCA1/2 Zygosity

In the TCGA-PRAD cohort, systemic therapy exposure was uncommon, and overall survival events were infrequent, with 26 deaths overall and two deaths each in the monoallelic and biallelic strata **(Table 1)**. Under these conditions, therapy class-specific Cox models with therapy-by-zygosity interaction terms yielded unstable estimates with wide confidence intervals, and therefore, TCGA-PRAD therapy-stratified estimates are not emphasized.

In the SU2C/PCF cohort, systemic therapy uptake was high, and death events were observed across zygosity strata (Table 2). We evaluated associations between therapy class and overall survival using Cox proportional hazards models parameterized with therapy exposure (ever vs never), patient-level BRCA1/2 zygosity indicators (monoallelic and biallelic, with wild-type as reference), and therapy-by-zygosity interaction terms. **Figure 2 and Supplementary Information 1 Table S3** summarizes exposure versus non-exposure hazard ratios within each zygosity stratum, along with a joint Wald test p-value for interaction.

**Figure 2.**
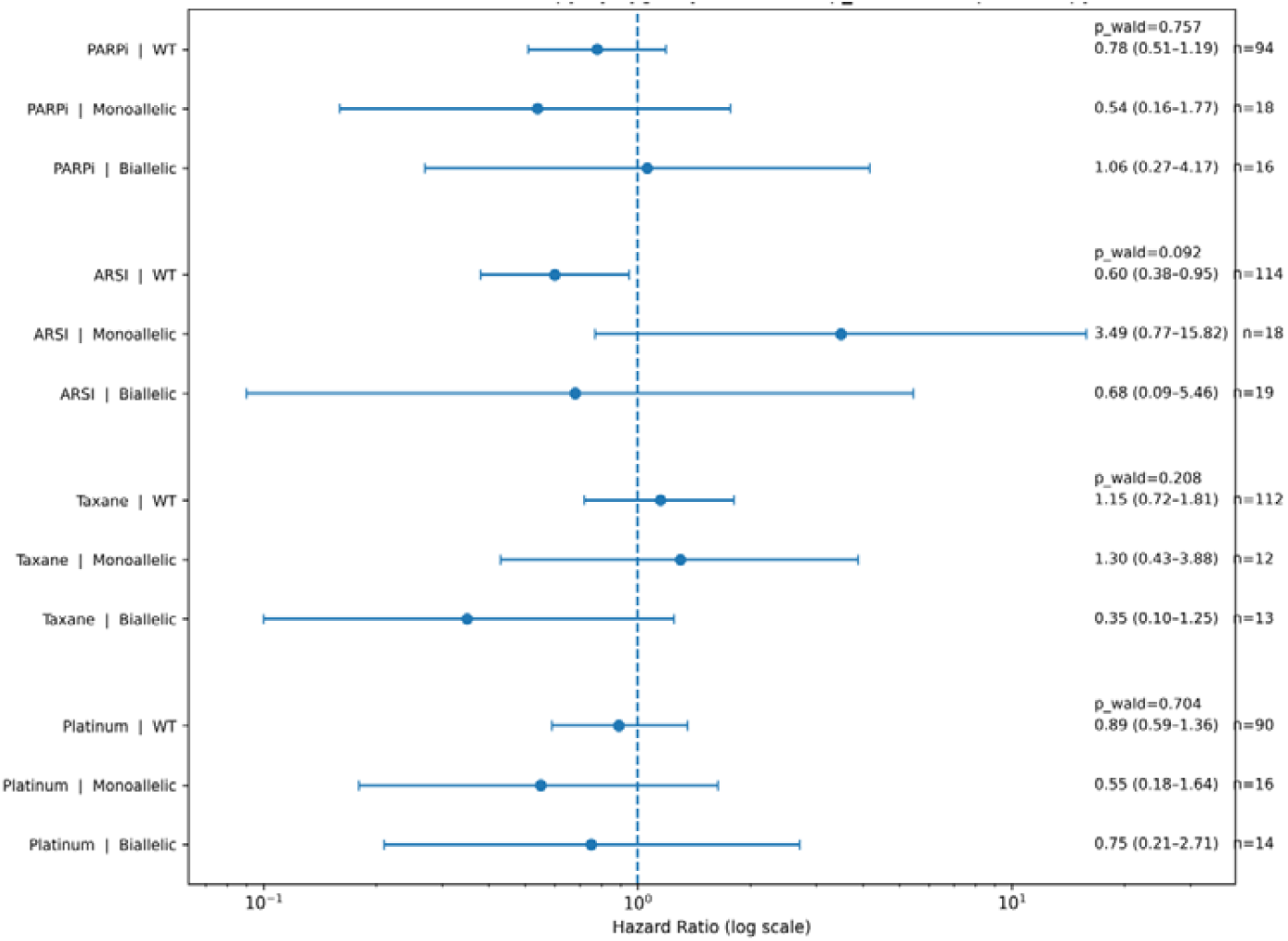
Systemic therapy class associations with overall survival by BRCA1/2 zygosity in SU2C/PCF. Forest plot of hazard ratios (HRs) for overall survival comparing ever-exposed versus never-exposed patients for each systemic therapy class, stratified by patient-level BRCA1/2 zygosity (wild-type, monoallelic, biallelic). Points indicate HRs from Cox proportional hazards models and horizontal bars indicate 95% confidence intervals. The number shown for each row denotes the number of exposed patients (n) within the corresponding zygosity stratum. For each therapy class, the reported p wald corresponds to a joint Wald test of therapy-by-zygosity interaction terms (monoallelic and biallelic, with wild-type as the reference stratum).

ARSI exposure was associated with improved overall survival within the WT stratum (HR 0.60, 95% CI 0.38 to 0.95). The corresponding estimates in altered strata were imprecise, with a higher hazard estimate in the monoallelic group (HR 3.49, 95% CI 0.77 to 15.82) and a lower hazard estimate in the biallelic group (HR 0.68, 95% CI 0.09 to 5.46). The interaction test for ARSI suggested a possible, but not definitive, departure from homogeneity across zygosity strata (interaction p = 0.092). For PARP inhibitors, the association estimates were directionally favorable in WT (HR 0.78, 95% CI 0.51 to 1.19) and monoallelic disease (HR 0.54, 95% CI 0.16 to 1.77), but not in biallelic disease (HR 1.06, 95% CI 0.27 to 4.17), and there was no evidence of effect modification (interaction p = 0.757). For taxanes, estimates were close to null in WT (HR 1.15, 95% CI 0.72 to 1.81) and monoallelic disease (HR 1.30, 95% CI 0.43 to 3.88), while the biallelic estimate suggested a lower hazard (HR 0.35, 95% CI 0.10 to 1.25), but the interaction test did not support heterogeneity (interaction p = 0.208). For platinum, estimates were broadly similar across strata (WT HR 0.89, 95% CI 0.59 to 1.36; monoallelic HR 0.55, 95% CI 0.18 to 1.64; biallelic HR 0.75, 95% CI 0.21 to 2.71) with no evidence of effect modification (interaction p = 0.704).

## DISCUSSION

This multicohort, desk-based analysis evaluated BRCA1/2 zygosity as a prognostic and potentially predictive feature across primary (TCGA-PRAD) and metastatic (SU2C/PCF) prostate cancer. BRCA1/2 alterations were uncommon in the TCGA-PRAD and more frequent in SU2C/PCF with BRCA2 predominating. Biallelic inactivation was infrequent in both cohorts. In TCGA-PRAD, overall survival showed only modest separation across wild-type, monoallelic, and biallelic strata and did not reach statistical significance, whereas in SU2C/PCF metastatic disease both altered strata had shorter median survival than wild-type with a significant global log-rank test. Exploratory treatment-class analyses did not demonstrate consistent or statistically robust effect modification by zygosity for androgen receptor signaling inhibitors (ARSI), PARP inhibitors, taxanes, or platinum agents. Collectively, these findings suggest that BRCA1/2 zygosity conveys prognostic information in metastatic disease in this dataset, while evidence for prognostic separation in primary disease remains limited.

In TCGA-PRAD, the small number of monoallelic and biallelic cases and the low number of survival events constrained statistical power. Median overall survival was numerically shortest in biallelic disease (55.0 months) compared with monoallelic (78.0 months) and wild-type (80.0 months), but confidence intervals overlapped and the global comparison was not significant. Accordingly, the primary cohort should be interpreted as underpowered to confirm or exclude a prognostic role of zygosity in localized disease rather than as evidence of prognostic neutrality. In contrast, SU2C/PCF included more BRCA1/2-altered cases and demonstrated clearer separation: median overall survival was 22.0 months (wild-type), 14.0 months (monoallelic), and 16.0 months (biallelic), consistent with reports that BRCA-deficient or HRR-altered mCRPC behaves more aggressively than HRR-intact disease [3,5,6]. The slightly shorter survival in monoallelic versus biallelic disease likely reflects heterogeneity within the monoallelic category (including cryptic second hits) and residual misclassification from reliance on public mutation, copy-number, and LOH exports, compounded by modest altered-stratum sizes.

Biologically, multiple lines of evidence support that biallelic BRCA1/2 inactivation and functional HRR deficiency are most closely aligned with PARP inhibitor synthetic lethality [11,12]. In TOPARP-B, outcomes under PARP inhibition were particularly favorable in tumors with BRCA2 homozygous loss, whereas monoallelic alterations and non-BRCA HRR alterations showed more variable benefit [13]. Functional RAD51-based assays further demonstrate that nominal genomic HRR alterations do not invariably translate into functional HRR deficiency and can stratify tumors that share similar genomic labels [14]. In this context, our zygosity-aware operationalization (including allowance for dual BRCA1/2 configurations) extends these concepts to a clinicogenomic setting; the predominance of BRCA2-only alterations is consistent with prostate cancer HRR epidemiology [3,12] and supports the value of refined zygosity calls for BRCA2-driven metastatic disease.

For treatment associations in SU2C/PCF, ever/never exposure models with joint interaction testing suggested limited evidence of zygosity-based effect modification. ARSI exposure was associated with improved survival in wild-type disease (HR 0.60; 95% CI 0.38–0.95), while estimates in altered strata were imprecise and the interaction test was at most suggestive (p=0.092). PARP inhibitor estimates were directionally favorable in wild-type and monoallelic disease, near null in biallelic disease, and showed no interaction (p=0.757); taxane and platinum associations were broadly similar across strata. These observational findings should be interpreted alongside randomized evidence demonstrating PARP inhibitor activity in molecularly selected mCRPC (TRITON3) and improved progression outcomes with PARP inhibitor combinations (PROpel, MAGNITUDE, TALAPRO-2) [7,8,9,10]. Differences in data structure likely limited detectability of effect modification in SU2C/PCF—exposure was not time-aligned, treatment sequencing was not modeled, and covariate adjustment was constrained—leaving susceptibility to confounding by indication, immortal-time bias, and selection effects. Strengths of this work include separate analysis of primary versus metastatic disease, prespecified two-hit zygosity definitions without pathogenicity reinterpretation, and reproducibility using public datasets. Key limitations include low event counts in TCGA-PRAD, potential monoallelic/biallelic misclassification, heterogeneous index dates, and observational therapy definitions without time-varying modeling; future studies with richer timelines, harmonized CN/LOH calling, and integration of functional HRD assays are needed to better link genomic zygosity, functional loss, and outcomes across therapy classes [6,11,13,14].

## CONCLUSION

This multicohort analysis indicates that BRCA1/2 zygosity is associated with overall survival in metastatic but not clearly in primary prostate cancer within the constraints of available data. In SU2C/PCF, both monoallelic and biallelic alterations corresponded to shorter survival than wild-type, supporting the adverse prognostic role of BRCA1/2 alteration in advanced disease. In TCGA-PRAD, limited events reduced power to detect differences. Exploratory, ever-exposed treatment models did not demonstrate consistent effect modification by zygosity across major systemic therapy classes and should not be viewed as comparative effectiveness estimates. Collectively, these findings support zygosity-aware annotation as a feasible and informative descriptor for metastatic prostate cancer in large genomic datasets, while underscoring the need for validation in cohorts with richer clinical annotation, standardized time origins, time-dependent treatment modeling, and integration of functional homologous recombination repair deficiency assays.

## Data Availability

All data produced in the present study are available upon reasonable request to the authors

https://gdc.cancer.gov/

https://www.cbioportal.org/

## Acknowledgements

The authors acknowledge the TCGA Research Network and the NCI Genomic Data Commons for access to the TCGA-PRAD dataset, and the SU2C/PCF Dream Team and cBioPortal for making metastatic prostate cancer data publicly available

## Authors’ Contribution

All authors contributed to the conception and design of the study, data acquisition, data analysis and interpretation, drafting of the manuscript, critical revision, and approval of the final manuscript.

## REFERENCES

1. Bray F, Laversanne M, Sung H, Ferlay J, Siegel RL, Soerjomataram I. Global cancer statistics 2022: GLOBOCAN estimates of incidence and mortality worldwide for 36 cancers in 185 countries. CA Cancer J Clin. 2024;74:229–263. doi:10.3322/caac.21834.

2. Schaeffer EM, Srinivas S, Adra N, An Y, Barocas D, Bitting R, et al. Prostate cancer, version 4.2023, NCCN clinical practice guidelines in oncology. J Natl Compr Canc Netw. 2023;21(10):1067–1096. doi:10.6004/jnccn.2023.0050.

3. Shui IM, Burcu M, Shao C, Chen C, Liao CY, Jiang S, et al. Real world prevalence of homologous recombination repair mutations in advanced prostate cancer: An analysis of two clinico genomic databases. Prostate Cancer Prostatic Dis. 2024;27:728–735. doi:10.1038/s41391-023-00764-1.

4. Yu EY, Rumble RB, Agarwal N, Cheng HH, Eggener SE, Bitting RL, et al. Germline and somatic genomic testing for metastatic prostate cancer: ASCO guideline. J Clin Oncol. 2025;43(6):748–758. doi:10.1200/JCO-24-02608.

5. Fettke H, Dai C, Kwan EM, Zheng T, Du P, Ng N, et al. BRCA deficient metastatic prostate cancer has an adverse prognosis and distinct genomic phenotype. EBioMedicine. 2023;95:104738. doi:10.1016/j.ebiom.2023.104738.

6. Olmos D, Lorente D, Alameda D, Cattrini C, Romero Laorden N, Lozano R, et al. Treatment patterns and outcomes in metastatic castration resistant prostate cancer patients with and without somatic or germline alterations in homologous recombination repair genes. Ann Oncol. 2024;35(5):458–472. doi:10.1016/j.annonc.2024.01.011.

7. Clarke NW, Armstrong AJ, Thiery Vuillemin A, Oya M, Shore N, Loredo E, et al. Abiraterone and olaparib for metastatic castration resistant prostate cancer. NEJM Evid. 2022;1(9):EVIDoa2200043. doi:10.1056/EVIDoa2200043.

8. Chi KN, Rathkopf DE, Smith MR, Efstathiou E, Attard G, Olmos D, et al. Niraparib and abiraterone acetate for metastatic castration resistant prostate cancer. J Clin Oncol. 2023;41(18):3339–3351. doi:10.1200/JCO.22.01649.

9. Agarwal N, Azad AA, Carles J, Fay AP, Matsubara N, Heinrich D, et al. Talazoparib plus enzalutamide in men with first line metastatic castration resistant prostate cancer (TALAPRO 2): A randomised, placebo controlled, phase 3 trial. Lancet. 2023;402(10398):291–303. doi:10.1016/S0140-6736(23)01055-3.

10. Fizazi K, Piulats JM, Reaume MN, Ostler P, McDermott R, Gingerich JR, et al. Rucaparib or physician’s choice in metastatic prostate cancer. N Engl J Med. 2023;388(8):719–732. doi:10.1056/NEJMoa2214676.

11. Serritella AV, Taylor A, Haffner MC, Abida W, Bryce A, Karsh LI, et al. Therapeutic implications of homologous repair deficiency testing in patients with prostate cancer (Part 2 of 2). Prostate Cancer Prostatic Dis. 2025;28(3):601–609. doi:10.1038/s41391-024-00887-z.

12. Teyssonneau D, Margot H, Cabart M, Anonnay M, Sargos P, Corn PG. Prostate cancer and PARP inhibitors: Progress and challenges. J Hematol Oncol. 2021;14(1):51. doi:10.1186/s13045-021-01061-x.

13. Carreira S, Porta N, Arce Gallego S, Seed G, Llop Guevara A, Bianchini D, et al. Biomarkers associating with PARP inhibitor benefit in prostate cancer in the TOPARP B trial. Cancer Discov. 2021;11(11):2812–2827. doi:10.1158/2159-8290.CD-21-0007.

14. Arce Gallego S, Cresta Morgado P, Delgado Serrano L, Simonetti S, Mateo J, et al. Homologous recombination repair status in metastatic prostate cancer by next generation sequencing and functional immunofluorescence. Cell Rep Med. 2025;6(2):101937. doi:10.1016/j.xcrm.2025.101937.

