## Supplementary Information 1 for "Prognostic and Therapeutic Relevance of BRCA1/2 Zygosity in Prostate Cancer: A Multicohort Desk-Based Analysis"

**Table S1. Overall survival by BRCA1/2 zygosity in TCGA-PRAD (primary disease cohort, n = 300)**

| **Zygosity group** | **n** | **Median OS, months (95% CI)** | **OS at 24 months** | **OS at 60 months** | **OS at 96 months** |
| --- | --- | --- | --- | --- | --- |
| Wild-type | 272 | 80.0 (72.0–92.0) | 0.87 (0.83–0.91) | 0.60 (0.54–0.66) | 0.32 (0.26–0.39) |
| Monoallelic | 18 | 78.0 (56.0–>120.0) | 0.86 (0.70–0.96) | 0.58 (0.38–0.77) | 0.28 (0.11–0.51) |
| Biallelic | 10 | 55.0 (30.0–>120.0) | 0.78 (0.52–0.93) | 0.40 (0.18–0.65) | 0.20 (0.05–0.47) |

Global log-rank p value (WT vs monoallelic vs biallelic): p = 0.45

#### Table S2. Overall survival by BRCA1/2 zygosity in SU2C/PCF (metastatic disease cohort, n = 200)

| **Zygosity group** | **n** | **Median OS, months (95% CI)** | **OS at 6 months** | **OS at 12 months** | **OS at 24 months** | **OS at 36 months** |
| --- | --- | --- | --- | --- | --- | --- |
| Wild-type | 153 | 22.0 (19.0–25.0) | 0.83 (0.77–0.88) | 0.70 (0.63–0.76) | 0.42 (0.35–0.49) | 0.24 (0.18–0.31) |
| Monoallelic | 25 | 14.0 (10.0–19.0) | 0.72 (0.51–0.86) | 0.50 (0.30–0.68) | 0.32 (0.15–0.50) | 0.18 (0.06–0.35) |
| Biallelic | 22 | 16.0 (12.0–23.0) | 0.76 (0.55–0.89) | 0.56 (0.35–0.73) | 0.37 (0.19–0.55) | 0.20 (0.08–0.38) |

Global log-rank p value (WT vs monoallelic vs biallelic): **p = 0.04**

#### Table S3. Therapy exposure and overall survival within BRCA1/2 zygosity strata in SU2C/PC

| **Systemic Therapy** | **Wild-type HR** | **Monoallelic HR** | **Biallelic HR** | **Interaction p** |
| --- | --- | --- | --- | --- |
| PARPi | 0.78 (0.51–1.19) | 0.54 (0.16–1.77) | 1.06 (0.27–4.17) | 0.757 |
| ARSI | 0.60 (0.38–0.95) | 3.49 (0.77–15.82) | 0.68 (0.09–5.46) | 0.092 |
| Taxane | 1.15 (0.72–1.81) | 1.30 (0.43–3.88) | 0.35 (0.10–1.25) | 0.208 |
| Platinum | 0.89 (0.59–1.36) | 0.55 (0.18–1.64) | 0.75 (0.21–2.71) | 0.704 |
